# Sustained Minimal Residual Disease Negativity in Multiple Myeloma is Associated with Stool Butyrate and Healthier Plant-Based Diets

**DOI:** 10.1101/2022.03.29.22272361

**Authors:** Urvi A. Shah, Kylee H. Maclachlan, Andriy Derkach, Meghan Salcedo, Kelly Barnett, Julia Caple, Jenna Blaslov, Linh Tran, Amanda Ciardiello, Miranda Burge, Tala Shekarkhand, Peter Adintori, Justin Cross, Matthew J. Pianko, Sham Mailankody, Neha Korde, Malin Hultcrantz, Hani Hassoun, Carlyn Tan, Sydney Lu, Dhwani Patel, Benjamin Diamond, Gunjan Shah, Michael Scordo, Oscar Lahoud, David J. Chung, Heather Landau, Saad Usmani, Sergio Giralt, Ying Taur, C. Ola Landgren, Gladys Block, Torin Block, Jonathan U. Peled, Marcel RM van den Brink, Alexander M. Lesokhin

## Abstract

Sustained minimal residual disease (MRD) negativity is associated with long-term survival in multiple myeloma (MM). The gut microbiome is affected by diet, and in turn can modulate host immunity, for example through production of short-chain fatty acids including butyrate. We examined the relationship of dietary factors, stool metabolites, and microbial diversity with sustained MRD negativity in patients on lenalidomide maintenance. At 3 months, higher stool butyrate concentration (p=0.037), butyrate producers (p=0.025) and α-diversity (p=0.0035) were associated with sustained MRD-negativity. Healthier dietary proteins, (from seafood and plants), correlated with butyrate at 3 months (p=0.009) and sustained MRD-negativity (p=0.05). Consumption of dietary flavonoids, plant nutrients with antioxidant effects, correlated with stool butyrate concentration (anthocyanidins p=0.01, flavones p=0.01, and flavanols p=0.02). This is the first study to demonstrate an association between a plant-based dietary pattern, stool butyrate production and sustained MRD-negativity in MM; providing rationale to evaluate a prospective dietary intervention.

**Statement of Significance:** We demonstrate an association between diet, the gut microbiome, and sustained MRD-negativity in MM. A healthy diet, with adequate plant and seafood protein, and containing flavonoids, associates with stool diversity, butyrate production and sustained MRD-negativity. These findings suggest dietary modification should be studied to enhance myeloma control.

**Key Points:**

1. In MM on lenalidomide maintenance, stool butyrate concentration at 3 months was associated with higher rates of MRD negativity at 12 months.
2. Increased seafood and plant proteins, dietary flavonoids, and diversity of dietary flavonoids correlated with stool butyrate concentrations.

## Introduction

Multiple myeloma (MM) remains incurable, however, sustained minimal residual disease (MRD) negativity following therapy represents the best predictor of survival.(1) Our prior studies in newly diagnosed MM demonstrated an increased relative abundance of *Eubacterium hallii* and *Faecalibacterium prausnitzii* in the stool samples collected following induction in MRD-negative patients.(2) In allogeneic hematopoietic cell transplantation, an increased abundance of *Eubacterium limosum* is associated with a lower risk of relapse and prolonged post-transplant survival.(3) These bacteria produce the short-chain fatty acid (SCFA) butyrate from dietary fiber and starch in plant foods. The SCFA butyrate can modulate systemic immunity through inhibition of NF-κB and histone deacetylases (HDAC), producing transcriptional modulation and a reduction in proinflammatory cytokines.(4-6)

Dietary composition plays a significant role in shaping the intestinal microbiome, with enrichment of stool butyrate concentration having been reported in individuals on a plant-based diet compared with an animal-based diet(7). In addition, dietary flavonoids, (plant derived nutrients) modulate both the microbiome and intestinal immune functions.(8, 9) We hypothesized that dietary factors that affect the microbiome, in particular the abundance of butyrate-producing bacteria or stool butyrate concentration, may be associated with MM outcomes. Here, in the context of lenalidomide maintenance therapy, we evaluated the relationship of dietary factors, stool metabolites and microbial composition with sustained MRD-negativity.

## Results

Baseline patient characteristics are presented in **Table 1**. Samples were available from 74 MM patients including 59 with assessment of habitual dietary patterns, and 49 with 16S sequencing of the stool microbiome. The overlap of dietary assessment and stool examination was present in 34 patients, of which 32 had stool butyrate concentration measurements (**Figure 1**). All patients had MRD status assessed at enrollment, with 42 being MRD-positive and 32 MRD-negative. Serial assessment of MRD status was performed in 68 patients at 12 months (m), 61 at 24m and 48 at 36m from enrollment. Sustained MRD-negativity was observed in 31 patients.

**Table 1.** Patient characteristics.

| Clinical variable | Value |
| --- | --- |
| Patients (n) | 74 |
| Gender |  |
| female, n (%) | 27 (36) |
| male, n (%) | 47 (64) |
| Age |  |
| ≤ 65 years, n (%) | 38 (51) |
| > 65 years, n (%) | 36 (49) |
| High-risk cytogenetics, n (%) | 21 (30) |
| Induction regimen |  |
| Dara-KRd | 4 (5) |
| KRd | 42 (57) |
| VRd | 18 (24) |
| CyBorD | 4 (5) |
| other | 6 (8) |
| Post-transplant status |  |
| yes, n (%) | 33 (45) |
| no, n (%) | 41 (55) |
| MRD-negativity at enrollment |  |
| yes, n (%) | 32 (43) |
| no, n (%) | 42 (57) |
| MRD-negativity at 12m |  |
| yes, n (%) | 31 (46) |
| no, n (%) | 37 (54) |
| MRD-negativity at 2y |  |
| yes, n (%) | 32 (52) |
| no, n (%) | 29 (48) |
| Sustained MRD-negativity, n (%) | 31 (46) |

High-risk cytogenetics % from 69 available results
MRD at 12m % from 68 available samples
MRD at 2y % from 61 available samples
Other includes Vd, KCyD, thal
Dara-KRd: Daratumumab, Carfilzomib, Lenalidomide, Dexamethasone
KRd: Carfilzomib, Lenalidomide, Dexamethasone
VRd: Bortezomib, Lenalidomide, Dexamethasone
CyBorD: Cyclophosphamide, Bortezomib, Dexamethasone
Vd: Bortezomib, Dexamethasone

**Figure 1.**
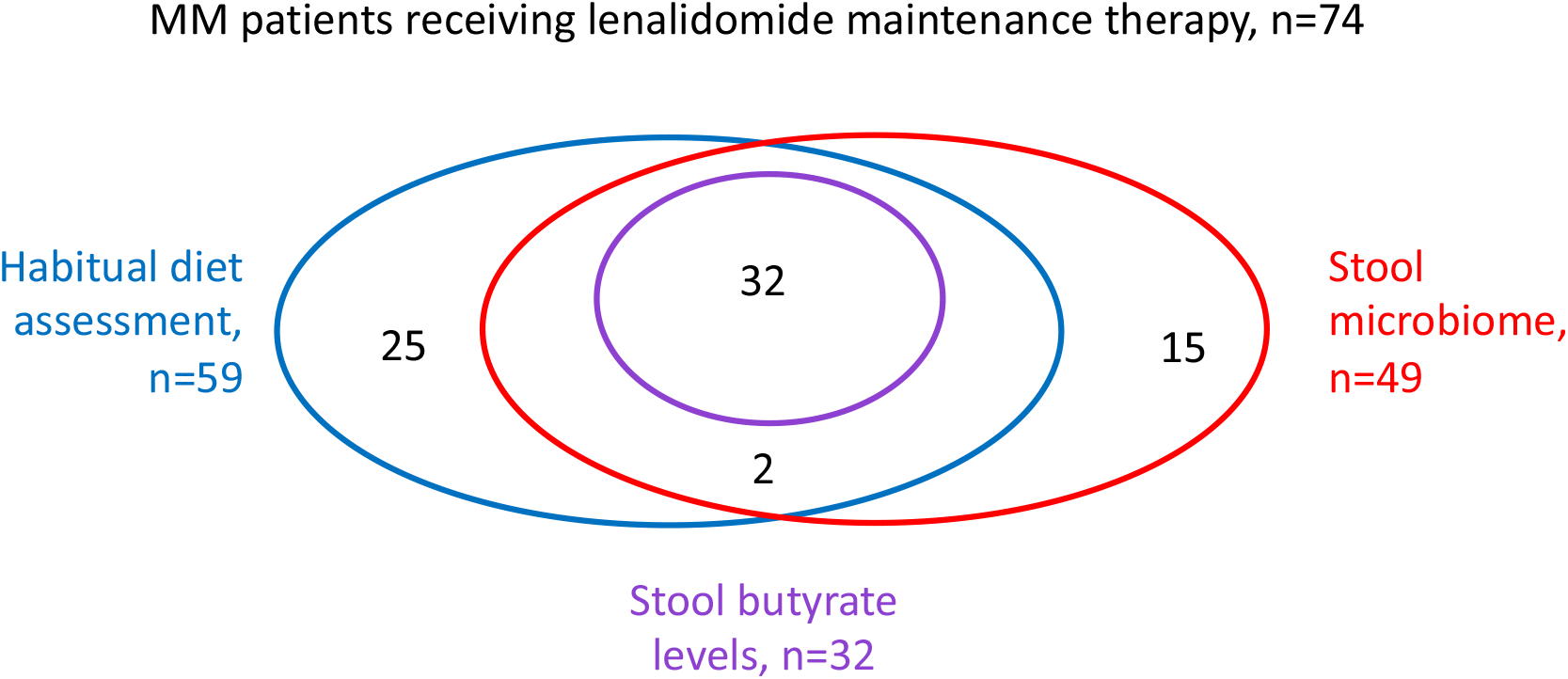
Patient samples.

As a subset of patients had undergone autologous hematopoietic cell transplantation (AHCT) prior to maintenance therapy (33/74, 45%), the timepoint for microbiome evaluation of 3m post-enrollment was chosen to allow resolution of post-AHCT reduction in microbiome diversity.(10) α-diversity of the fecal microbiome at 3m was significantly higher in those with sustained MRD-negativity (median 16.9, interquartile range [IQR] 14.8-28.0), compared with those without (median 11.9, IQR 9.9-18.6, p=0.0035) (**Figure 2a**). The relative abundance of predicted butyrate-producers was also significantly higher in patients with sustained MRD-negativity (median 0.093, IQR 0.072-0.100) than those without (median 0.054, IQR 0.035-0.094, p=0.025) (**Figure 2b**).

**Figure 2.**
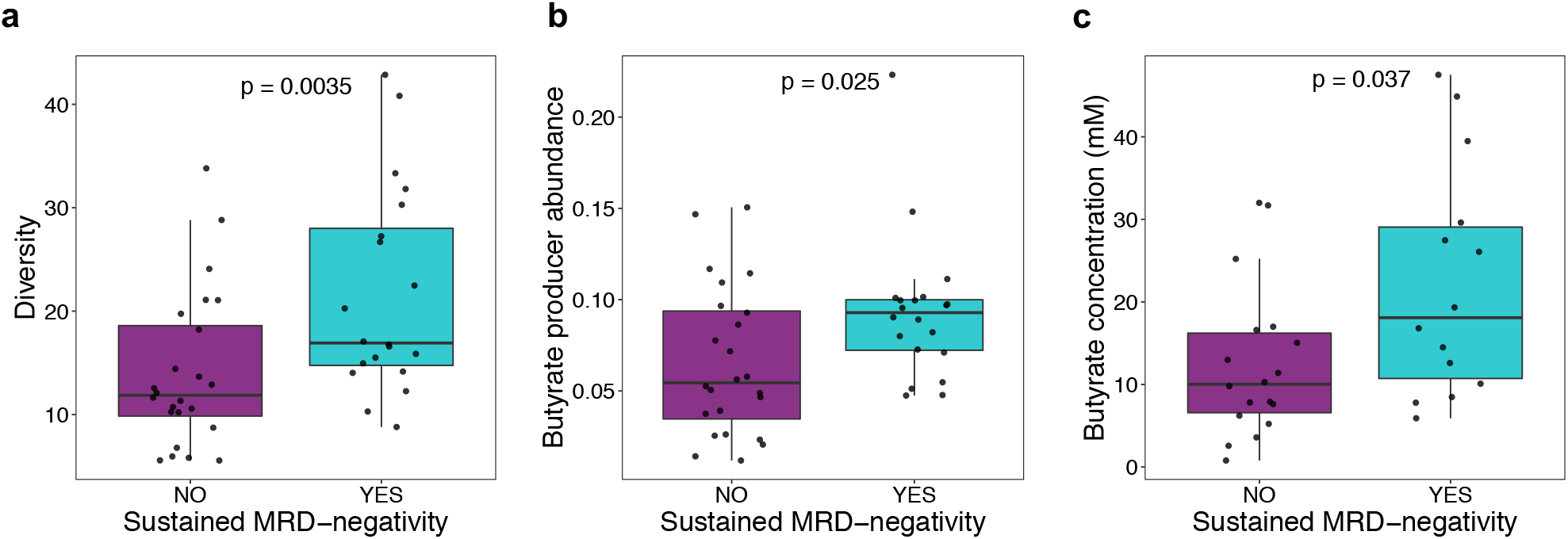
Stool α-diversity, abundance of butyrate producers and butyrate concentration associate with MRD-negativity. (a) Stool α-diversity at 3m by inverse Simpson index according to sustained MRD-status. (b) Relative abundance of butyrate producers at 3m according to sustained MRD-status. (c) Concentration of stool butyrate at 3m according to sustained MRD-status. (Achievement of sustained MRD negativity; Turquoise, yes; Purple, no).

Stool butyrate concentration at 3m was also significantly higher in patients who achieved sustained MRD-negativity (median 18.1, IQR 10.7-29.0mM) compared to those who did not (median 10.0mM, IQR 6.6-16.2mM) (p=0.037) (**Figure 2c**). These associations remained significant after adjusting for AHCT and high-risk cytogenetic features (deletion 1p32, gain 1q21, t(4;14) or deletion 17p). Other stool metabolites (acetate, propanoate, valerate, heptanoate, isobutyrate, methylbutyrate, isovalerate) did not correlate with MRD-status (p-values >0.1). Collectively, these data suggest that sustained MRD-negativity in MM is associated with higher α-diversity, relative abundance of butyrate producers and concentration of stool butyrate.

Considering that dietary factors play an important role in the production of SCFA, we examined the relationship between diet composition (components of HEI-2015 score), stool butyrate concentration, and subsequent MRD-status and identified certain significant correlations (**Supplementary Table 1**). The components total protein as well as seafood and plant protein were associated with stool butyrate concentration at 3m (R=0.5, p=0.004 and R=0.45, p=0.009 respectively). Seafood and plant proteins include seafood, nuts, seeds, soy products (excluding beverages), and legumes (beans and peas). The standard for maximum score is ≥0.8 cup equivalent per 1,000 kcal and standard for minimum score of zero is no seafood or plant proteins. These components also correlated with sustained MRD-negativity (p=0.01 and p=0.05 respectively). To further explore the evidence supporting a plant-based diet, we measured dietary flavonoids in this cohort. Total anthocyanidins (R=0.47, p=0.01), flavones (R=0.48, p=0.01), and flavanols (R=0.42, p=0.02) correlated with stool butyrate. Additionally, the Dietary Flavonoid Diversity Index was associated with butyrate concentration (R=0.46, p=0.008) and microbiome diversity (R=0.38, p=0.03) (**Supplementary Table 2**).

## Discussion

In the context of lenalidomide maintenance therapy for MM, we demonstrate for the first time an association between diet, the gut microbiome, and sustained MRD-negativity in MM. Our data show that sustained MRD-negativity among patients receiving lenalidomide is associated with higher microbial α-diversity, relative abundance of butyrate producers and concentration of stool butyrate measured after 3 months on lenalidomide maintenance. Together with our prior publication demonstrating increased butyrate producers, specifically *Eubacterium hallii* and *Faecalibacterium prausnitzii* in MRD-negative patients following induction therapy(2), this study further strengthens the hypothesis that specific microbiome features, especially butyrate levels may predict clinical outcomes in MM.

Butyrates have previously been shown to modulate immunity by exerting anti-inflammatory functions through inhibition of the transcription factor NF-κB, leading to reduced formation of proinflammatory cytokines.(4, 5) They also non-competitively inhibit HDACs, acting in the same way as panobinostat, an HDAC inhibitor with activity in MM.(4, 6)

The dietary associations described in our study are consistent with prior epidemiologic data. Higher HEI-2015 scores correlated with reduced cancer risk and mortality.(11, 12) In the EPIC Oxford study, including 61,647 individuals of which 65 developed MM, those on vegetarian and vegan diets had reduced risk of development of MM compared to meat eaters (relative risk 0.23; 95% confidence interval (CI) 0.09-0.59).(13) In the Nurses’ Health Study and Health Professionals Follow-up study that included 165,796 individuals with 423 MM cases and 345 deaths, those with healthier pre-diagnosis dietary pattern based on the alternative healthy eating index 2010 had lower MM mortality (hazard ratio 0.76; 95% CI 0.67-0.87).(14) The association of butyrate concentration with seafood and plant protein scores, dietary flavonoids, and dietary flavonoid diversity support the hypothesis that a diverse plant-based diet may have an impact on MM via butyrate production suggesting the potential for an underlying mechanistic basis.

Strengths of this study include the availability of simultaneous dietary, stool microbiome and metabolite assessment, as well as long term MRD data, and the consistent association between the relative abundance of predicted butyrate producers and stool butyrate levels. Limitations include the lack of untreated patient samples prior to myeloma therapy initiation, a small sample size that had all study assessments, dietary recall bias and potential confounding from other aspects of an individual’s lifestyle and circumstances.

Nevertheless, our data support the hypothesis that a healthy diet, with adequate high-quality plant and seafood protein, and dietary flavonoids may have a positive impact on stool diversity, butyrate production and MM outcomes (**Figure 3**).

**Figure 3.**
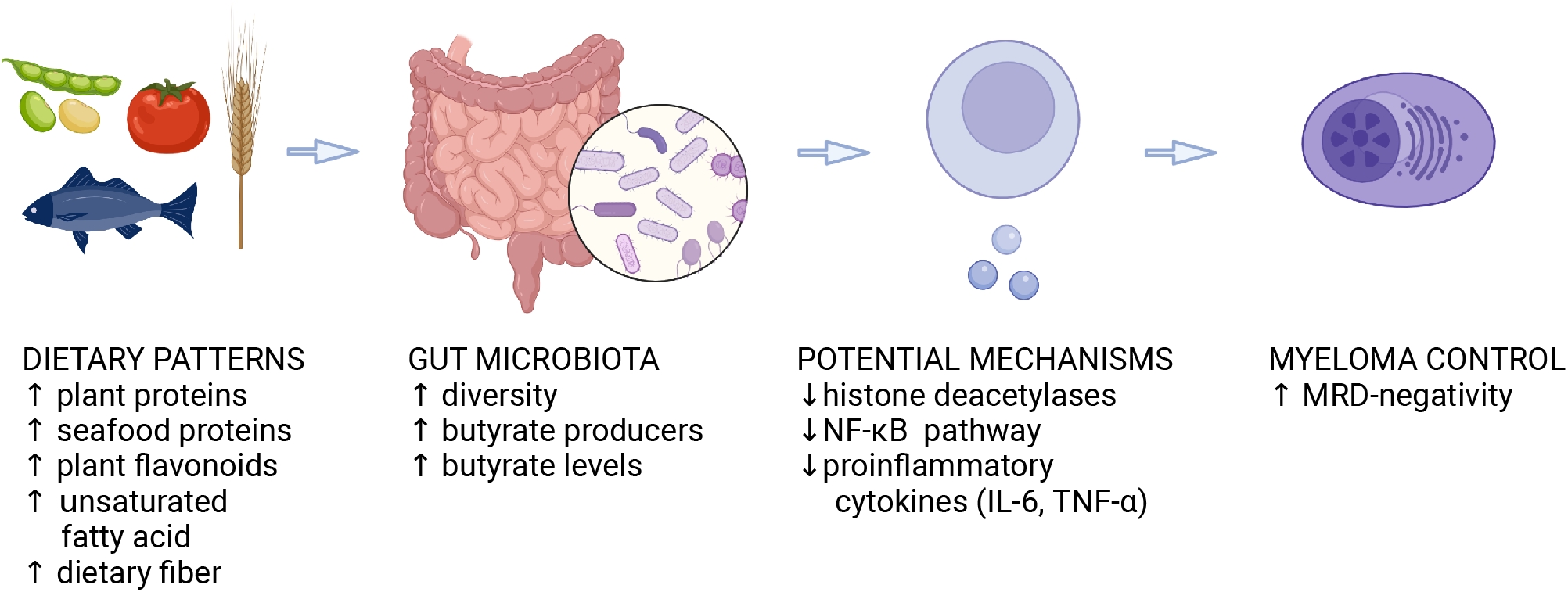
Schema for the hypothesis on how dietary composition may impact the intestinal microbiome and sustained MRD-negativity. Created with BioRender.com

In conclusion, our study suggests that lifestyle modification in the form of dietary change may potentially contribute to MM control. We therefore believe that further study of prospective dietary interventions in plasma cell disorders is warranted and have initiated a prospective whole-foods plant based dietary interventional trial in MM precursor disease (NCT04920084, NUTRIVENTION).

## Methods

MM patients eligible for maintenance during first-line therapy were enrolled prospectively, and received lenalidomide for up to 5 years.(15) MRD status was evaluated at enrollment then annually using a validated bone marrow–based flow cytometric assay^(16)^ with a sensitivity of at least 10^−5^. Treatment responses were assessed according to International Myeloma Working Group consensus criteria(17), with sustained MRD-negativity defined as MRD-negativity at two consecutive time points 1 year apart between enrolment, 12m and 24m. Progression-free survival was not used as an endpoint given the low rate of clinical progression.

Stool samples were collected and analyzed as detailed previously(2) with identification of predicted butyrate-producing bacteria.(18) The relative abundance of predicted butyrate-producers and microbiome α-diversity were calculated from 16S microbiome profiles in samples collected 3 months (m) from enrollment. We performed direct quantitation of stool metabolite concentrations using gas chromatography–mass spectrometry on the same stool samples.

Comparison of α-diversity (as measured by the inverse Simpson index) and relative abundance of butyrate producers were assessed using the Wilcoxon rank sum test. The association between dietary and microbiome data were evaluated by Spearman’s rank correlation coefficient (R). The Kruskal–Wallis test was used to evaluate association between sustained MRD-negativity, microbiome, and dietary measurements. In this exploratory analysis we declared statistical significance at a two-sided significance level below 0.05.

Habitual dietary patterns were collected using the Block Food Frequency Questionnaire 2014 (FFQ)(19) and summarized using the United States Department of Agriculture’s (USDA) Healthy Eating Index 2015 score (HEI-2015)(20) and a newly developed Dietary Flavonoid Diversity Index (*DFDI*) by NutritionQuest. The HEI-2015 is a measure of diet quality, assessing 13 nutrient and food group components, with higher scores indicating healthier diets.(20) Flavonoid nutrient values were calculated from the FFQ based on USDA data. The DFDI measures the diversity of flavonoid intake from foods and beverages consumed at least once per week, following the Berry-Index method(21, 22); scores range from 0-1, with higher scores indicating a greater diversity of flavonoid intake.

## Data Availability

All data produced in the present work are contained in the manuscript.

## Acknowledgements

This study is funded in part through the NIH/NCI Cancer Center Support Grant P30 CA008748, Sawiris Family Fund, Paula and Rodger Riney Foundation and Celgene.

U.A.S. received research support from the American Society of Hematology Clinical Research Training Institute, Parker Institute for Cancer Immunotherapy, International Myeloma Society, Paula and Rodger Riney Foundation, TREC Training Workshop R25CA203650 (PI: Melinda Irwin), NCI MSK Paul Calabresi Career Development Award for Clinical Oncology K12 CA184746, HealthTree Foundation and the Allen Foundation Inc.

K.H.M received research support from a Multiple Myeloma Research Foundation Fellow Award, an American Society of Hematology Restart Research Award and a Royal Australasian College of Physicians Research Establishment Grant.

MJP received research support from National Institutes of Health grants (National Institute of Aging: P30-AG024824; National Cancer Institute: P30-CA046592, and the National Center for Advancing Translational Sciences award UL1-TR00457.

AML received research support from NCI 1R01CA249981-01, Sawiris Family Fund and Paula and Rodger Riney Foundation.

JUP reports funding from NHLBI NIH Award K08HL143189, the MSKCC Cancer Center Core Grant NCI P30 CA008748. This research was supported by the Parker Institute for Cancer Immunotherapy at Memorial Sloan Kettering Cancer Center; J.U.P is a member of the Parker Institute for Cancer Immunotherapy.

MVDB was supported by National Cancer Institute award numbers, R01-CA228358, R01-CA228308, and P01-CA023766; National Heart, Lung, and Blood Institute (NHLBI) award number R01-HL123340 and R01-HL147584; National Institute of Aging award number P01-AG052359; Starr Cancer Consortium, and Tri Institutional Stem Cell Initiative. Additional funding was received from The Lymphoma Foundation, The Susan and Peter Solomon Divisional Genomics Program, Cycle for Survival, and the Parker Institute for Cancer Immunotherapy.

AML received research support from NCI 1R01CA249981-01, Sawiris Family Fund and Paula and Rodger Riney Foundation.

## Authorship Contributions

UAS and AML conceived and designed the study.

UAS, KHM, AD, AML analyzed, verified, and interpreted the data

MS, KW, JC, JB, LT, AC, MB, TS, PA, MJP, SM, NK, MH, HH, CT, BD, SL, DP, GS, MS, OLah, DC, HL, SU, OLan, SG collected and assembled the data.

JC, YT, JUP, MVDB, UAS, AML, AD, KHM analyzed and interpreted the microbiome data.

GB, TB, UAS, AML, AD, KHM analyzed and interpreted the nutrition data.

UAS, KHM, AML had full access to the data and share final responsibility for submission of the publication.

All authors wrote and approved of the article and are accountable for publication.

## Disclosure of Conflicts of Interest

U.A.S. has received grants and research support from Celgene/Bristol Myers Squibb, Janssen paid to the institution, personal fees from Janssen, all outside of the submitted work.

SG reports personal fees and advisory role (scientific advisory board) from Actinnum, Celgene, Bristol Myers Squibb, Sanofi, Amgen, Pfizer, GlaxoSmithKline, JAZZ, Janssen, Omeros, Takeda, and Kite, outside the submitted work.

HH reports grants from Celgene, during the conduct of the study; and grants from Celgene, Takeda, and Janssen, outside the submitted work.

NK reports research funding through Amgen and participates in advisory board with Medimmune.

OLah reports serving on Advisory Board for MorphoSys.

OLan reports grants from Amgen, Janssen, and Takeda; Data Monitoring Committee from Janssen, Merck, and Takeda; and personal fees from Amgen, Janssen, GlaxoSmithKline, AstraZeneca, and The Binding Site, outside the submitted work.

AML reports grants from Novartis, during the conduct of the study; grants from Bristol Myers Squibb; personal fees from Trillium Therapeutics; grants, personal fees and non-financial support from Pfizer; and grants and personal fees from Janssen, outside the submitted work. AML also has a patent US20150037346A1 with royalties paid.

SM reports research funding from Allogene Therapeutics, Juno/Bristol Myers Squibb, Takeda Oncology, and Janssen Oncology; personal fees from Plexus communication, and Physician Education Resource, outside the submitted work.

MS reports personal fees and research funding from Angiocrine Bioscience and Omeros Corporation; personal fees from McKinsey & Company, Kite – A Gilead Company, and i3Health, outside the submitted work.

GLS reports research funding from Janssen and Amgen outside the submitted work.

MJP reports research funding from Trillium Therapeutics, Nektar Therapeutics, and Regeneron Pharmaceuticals; personal fees from Karyopharm, Oncopeptides, Takeda, Janssen, and Curio Science, MJH Life Sciences, outside the submitted work.

CT reports other from Janssen Research and Development outside the submitted work.

BD reports honoraria from Medscape and Sanofi outside the submitted work.

SZU reports grant support from Amgen, Array Biopharma, BMS, Celgene, GSK, Janssen, Merck, Pharmacyclics, Sanofi, Seattle Genetics, SkylineDX and Takeda; personal fees from Amgen, BMS, Celgene, EdoPharma, Genentech, Gilead, GSK, Janssen, Oncopeptides, Sanofi, Seattle Genetics, SecuraBio, SkylineDX, Takeda and TeneoBio; outside the submitted work.

JUP reports research funding, intellectual property fees, and travel reimbursement from Seres Therapeutics, and consulting fees from DaVolterra, CSL Behring, and from MaaT Pharma. He serves on an Advisory board of and holds equity in Postbiotics Plus Research. He has filed intellectual property applications related to the microbiome (reference numbers #62/843,849, #62/977,908, and #15/756,845).

MVDB has received research support and stock options from Seres Therapeutics and stock options from Notch Therapeutics and Pluto Therapeutics; he has received royalties from Wolters Kluwer; has consulted, received honorarium from or participated in advisory boards for Seres Therapeutics, WindMIL Therapeutics, Rheos Medicines, Merck & Co, Inc., Magenta Therapeutics, Frazier Healthcare Partners, Nektar Therapeutics, Notch Therapeutics, Forty Seven Inc., Ceramedix, Lygenesis, Pluto Therapeutics, GlaskoSmithKline, Da Volterra, Novartis (Spouse), Synthekine (Spouse), and Beigene (Spouse); he has IP Licensing with Seres Therapeutics and Juno Therapeutics; and holds a fiduciary role on the Foundation Board of DKMS (a nonprofit organization).

The remaining authors declare no potential conflicts of interest.

