## Supplemental Data for "Sustained Minimal Residual Disease Negativity in Multiple Myeloma is Associated with Stool Butyrate and Healthier Plant-Based Diets"

**Supplementary Table 1. Association of HEI2015 dietary features with butyrate concentration and sustained MRD-negativity.**

| | **Butyrate level at 3m** | | **Sustained MRD-negativity** |
|---|---|---|---|
| | R | p-value | p-value |
| **Total Vegetables** | 0.24 | 0.182 | 0.32 |
| **Greens and Beans** | 0.25 | 0.16 | 0.64 |
| **Total Fruit** | 0.01 | 0.969 | 0.36 |
| **Whole Fruit** | 0.17 | 0.357 | 0.47 |
| **Whole Grain** | -0.03 | 0.874 | 0.22 |
| **Total Dairy** | -0.3 | 0.096 | 0.75 |
| **Total Protein** | 0.5 | 0.004 | 0.01 |
| **Seafood and Plant Protein** | 0.45 | 0.009 | 0.05 |
| **Fatty Acids** | 0.27 | 0.14 | 0.95 |
| **Sodium** | 0.08 | 0.673 | 0.77 |
| **Refined Grain** | 0.26 | 0.148 | 0.25 |
| **Added Sugar** | 0.27 | 0.132 | 0.16 |
| **Saturated Fatty Acid** | 0.13 | 0.468 | 0.22 |
| **Total HEI-2015 score** | 0.25 | 0.17 | 0.36 |

**Supplementary Table 2. Association of dietary flavonoids with butyrate concentration.**

| Dietary Flavonoids | Butyrate concentration at 3m | |
|---|---|---|
| | R | p-value |
| **Total anthocyanidins** | 0.47 | 0.01 |
| **Total flavan-3-ols** | 0.31 | 0.08 |
| **Total flavanones** | -0.12 | 0.05 |
| **Total flavones** | 0.48 | 0.01 |
| **Total flavanols** | 0.42 | 0.02 |
| **Total isoflavones** | 0.16 | 0.37 |
| **Total flavonoids** | 0.27 | 0.13 |
| **Dietary Flavonoid Diversity Index** | 0.46 | 0.008 |
